# Trends and Cross-Country Inequalities in Dengue, 1990-2021

**DOI:** 10.1101/2024.12.17.24319184

**Authors:** Mingzhu Zhou, Yong He, Liangmiao Wu, Kaiyuan Weng

**Author notes:** These authors contributed equally to this work (Mingzhu Zhou, Yong He).

## Abstract

**Background:** The existing body of literature is deficient in the most recent data regarding the global perspective of dengue fever and its associated health inequities.Our aim is to assess the global burden of dengue fever and its health inequities from 1990 to 2021.

**Methods:** We utilized the Global Burden of Disease (GBD) database for epidemiological trends, demographic analysis, and epidemiological decomposition. Cross-national inequality and predictive modeling for the global dengue burden up to 2051 were also performed.

**Results:** Globally, dengue fever incidence, prevalence, DALYs, and mortality showed increasing trends with significant international disparities. From 1990 to 2021, ASR for incidence and prevalence rose by 1.83% (95% CI: 1.58%-2.08%), and for DALYs and mortality by 1.33% (95% CI: 1.10%-1.57%) and 1.70% (95% CI: 1.45%-1.94%), respectively. Age-period-cohort-model analysis revealed a positive correlation between dengue fever incidence and age, with mortality sharply increasing in those over 80. Decomposition of dengue fever DALYs burden showed population growth as the main contributor to the global burden, with varying impacts across SDI quintiles. Inequalities in dengue fever burden related to SDI persisted, shifting benefits from impoverished to affluent populations. BAPC model projections suggest stabilization of incidence and prevalence ASRs, with declining DALYs and mortality ASRs, particularly for females.

**Conclusion:** This study elucidates the changes in the burden of dengue fever against the backdrop of a burgeoning global population, severe aging, and pronounced health inequities across nations, quantifying these alterations and forecasting the trends in the disease burden over the next three decades. Concurrently, the research proposes effective measures for various countries and regions to mitigate health inequities associated with dengue fever and to reduce the associated disease burden.

## Introduction

Dengue fever, an acute vector-borne infectious disease caused by the Dengue virus, poses a significant challenge to public health on a global scale [1]. This viral disease is primarily transmitted by the Aedes aegypti and Aedes albopictus mosquitoes, with approximately 390 million people infected annually, out of which 96 million cases exhibit clinical symptoms [2]. As global climate change and urbanization accelerate, the endemic regions of dengue fever are expanding, and the disease burden is increasing, particularly in tropical and subtropical areas [3].

From a virological perspective, the Dengue virus has four distinct serotypes, namely DENV-1, DENV-2, DENV-3, and DENV-4 [4], and exhibits a high mutation rate, with each genotype potentially forming its own quasi-special group [5]. Vaccine developers must address the challenge of ensuring that their candidate vaccines induce a strong neutralizing immune response against all serotypes without causing antibody-dependent enhancement (ADE) [6]. Regrettably, despite the availability of vaccines against Dengue fever [7], the implementation of vaccination in highly endemic regions must also consider the sero prevalence in the target population [8]. Most importantly, without early detection and proper medical care, DENV can lead to fatal outcomes such as Dengue fever (DF), Dengue hemorrhagic fever (DHF), and Dengue shock syndrome (DSS) [9], with the mortality rate for severe Dengue infections potentially exceeding 20% [10].

Due to the significant negative impact of dengue fever, existing studies have attempted to address the issue by identifying the determinants of dengue incidence [11, 12], predicting its incidence [13,14], and estimating the resulting economic burden [15,16]. However, they have overlooked the varying impacts of different public health policies across countries on the incidence, prevalence, DALYs, and even mortality rates of dengue fever among different age groups.

Understanding the geographical distribution and disease burden of dengue fever, as well as the public health policies of different countries, is crucial for determining how to optimally allocate limited resources available for dengue control and for assessing the international impact of such activities [17]. Previous research has indicated that the unequal distribution of disease burden places greater health and economic pressures on resource-poor areas and vulnerable populations. This inequality is not only evident between different countries and regions but also among different socioeconomic groups.

This study will employ a multidisciplinary approach, integrating epidemiological and public health policy research, to conduct an in-depth analysis of the trends and inequalities in the global disease burden of dengue fever. By collecting and analyzing global dengue case data, the aim is to reveal the characteristics of the global and regional distribution of the dengue disease burden, identify key factors contributing to disparities in disease burden, and explore potential strategies to reduce health inequalities. Through this research, we anticipate providing valuable information and recommendations to global health policymakers, in order to achieve more equitable and effective global health governance.

## Method

### Data Source

The GBD 2021 utilized the most recent epidemiological data and improved standardization methods to comprehensively assess health losses from 371 diseases, injuries, and risk factors across 204 countries and territories, stratified by age and sex[18,19]. GBD 2021 integrated various data sources, each with a unique identifier, listed in the Global Health Data Exchange (GHDx). The collected data were modeled using Spatio-Temporal Gaussian Process Regression (ST-GPR), allowing for the smoothing of age, time, and location in areas lacking complete datasets[8].

For epidemiological data with known biases, the Meta-regression-Bayesian (MR-BRT) model was employed to adjust for biases resulting from research methodologies. Incidence and prevalence were modeled using DisMod-MR 2.1 (Disease Modeling Meta-Regression; version 2.1), while Disability-Adjusted Life Years (DALYs) were estimated by location, age, sex, and year, and mortality was modeled using the Cause of Death Ensemble model (CODEm). In this study, estimates and their 95% uncertainty intervals (UI) were extracted from GBD 2021 for dengue fever’s incidence, prevalence, DALYs, and mortality[18]. All rates are reported per 100,000 people. Additionally, the attributable burden estimates were stratified by Socio-demographic Index (SDI) quintiles, representing a composite measure of income, education, and fertility conditions, quantifying the socio-demographic development level of a country or region, including five SDI quintiles (i.e., low, low-middle, middle, high-middle, high), representing five levels of development, and presented in counts, age-standardized rates, and rankings.

### Trend Analysis

The trend in Age-Standardized Rates (ASR) measured by the Annual Percentage Change (EAPC) is a more reliable indicator for monitoring changes in disease patterns[20]. A linear regression model was constructed as (y = *α* + *β* x + *є*), where (y = ln(ASR)), (x) represents the calendar year, and (*є*) is the error term, with (*β*) indicating the positive or negative trend in ASR. The EAPC was then calculated as ((exp(*β*) - 1) × 100%), and its 95% confidence interval (CI) was derived from the model[21]. If the EAPC estimate and the lower limit of its 95% CI are both >0, the ASR is considered to be increasing. Conversely, if the EAPC estimate and the upper limit of its 95% CI are both <0, the ASR is decreasing. Otherwise, the ASR is considered stable.

An age-period-cohort model studies the changes in a variable over time by simultaneously including three temporal dimensions to estimate the effects of age, period, and birth cohort on incidence and mortality rates. In this model, the age effect represents changes in the variable throughout an individual’s life, while the period effect represents the impact of environmental factors affecting the entire population. Additionally, the birth cohort effect refers to changes in the variable due to similar life events experienced by those born in the same year[22,23]. However, the inevitable identification issues arising from multicollinearity between ages, periods, and birth cohorts lead to challenges in estimating the effects of each. To address this, an Intrinsic Estimator (IE) method for age-period-cohort models is required[24,25], which uses principal component regression analysis to explain the variation effects on the three temporal trends and provides relatively efficient estimates. The IE method’s age-period-cohort model is based on the Poisson distribution. The data were also re-analyzed by continuous encoding into 5-year age groups (under 5, 5-10, …, 90-95, over 95), continuous 5-year periods (1992-1997, 1997-2002, …, 2017-2021), and corresponding continuous 5-year birth cohorts (1892-1897, 1897-1902, …, 2017-2021), and by comparing incidence and mortality in the same age group across different periods to estimate the net age, period, and birth cohort effects on dengue fever incidence and mortality rates. The IE method’s age-period-cohort model provides the effects of age, period, and birth cohort. These coefficients are then transformed into exponent values to determine the relative risks (RRs) of incidence and mortality at specific ages, periods, or birth cohorts compared to the average combined level across all ages, periods, or birth cohorts[26].

### Decomposition Analysis

To elucidate the contributing factors driving the changes in dengue fever DALYs from 1990 to 2021, we conducted a decomposition analysis stratified by sex, at the global and SDI quintile levels, accounting for demographic shifts, age structure, and epidemiological changes. Initially, DALYs for dengue fever were categorized into three subgroups based on gender: male, female, and all-gender. Subsequently, the DALYs rate, defined here as epidemiological change, was calculated as described by Xie et al.[27]: the number of DALYs at location, age, and year (DALYs_ay, py, ey_) can be computed as follows: 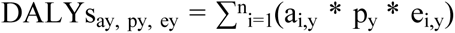, where DALYs a_y_, p_y_, e_y_ represents the DALYs influenced by age structure, population, and DALYs rate for year y; a_i,y_ is the population fraction for age group i of n groups in year y; p_y_ is the total population for year y; and e_i,y_ is the DALYs rate for age group i in year y. For instance, the impact of age structure is calculated as: [(DALY_a2021, p1990, e1990_ + DALY_a2021 p2021, e2021_)/3 + (DALY_a2021, p1990, e2021_ + DALY_a2021, p2021, e1990_)/6] - [(DALY_a1990, p2021, e2021_ + DALY_a1990, p1990, e1990_)/3 + (DALY_a1990, p2021, e1990_ + DALY_a1990, p1990, e2021_)/6]. The determination of their impact on population and epidemiological changes is identical.

### Cross-country Inequalities Analysis

Monitoring health inequalities can provide a foundation for evidence-based health planning, further improving policies, programs, and practices to reduce disparities in health distribution. In this study, we employed two standard indicators of absolute and relative gradients of inequality, namely the Slope Index of Inequality (SII) and the Concentration Index, to measure the inequality of dengue fever burden among countries and analyze the trends in global health inequalities from 1990 to 2021 [28,29]. The SII represents the health gap between the poorest and wealthiest groups, calculated by regressing the DALYs rates of all age groups in countries on a relative position scale associated with SDI. The relative position is determined by the midpoint of the population cumulative range sorted by SDI. A weighted regression model was used to account for heteroscedasticity. The Concentration Index was calculated by numerically integrating the area under the Lorenz concentration curve, fitted with the cumulative fraction of DALYs and the cumulative relative distribution sorted by SDI [30].

### Predictive Analysis

The aforementioned analyses focused on the burden of dengue fever over the past decades. To establish better public health policies and allocate health resources rationally, this study further predicted the burden of dengue fever over the next three decades. The Integrated Nested Laplace Approximation (INLA) with the Bayesian Age-Period-Cohort (BAPC) model offers better coverage and precision than the APC model. Therefore, this study utilized INLA with the BAPC model to approximate the marginal posterior distribution, forecasting the different burdens of dengue fever for males and females globally up to 2051 [31,32], avoiding several mixing and convergence issues associated with the Markov Chain Monte Carlo sampling techniques traditionally used for Bayesian methods [31].

All statistical analyses in this study were conducted using R version 4.3.2.

## Results

### Overall trends in Dengue burden using broad estimation method and descriptive analysis

Globally, from 1990 to 2021, the number of dengue fever cases, along with the Annual Incidence Rate (ASIR), Annual Prevalence Rate (ASPR), and DALYs, have all shown an overall increasing trend. Between 1990 and 2021, the ASIR and ASPR for dengue fever both rose by an average of 1.83% (95% CI: 1.58%-2.08%); the ASR for DALYs and mortality rates increased by 1.33% (95% CI: 1.10%-1.57%) and 1.70% (95% CI: 1.45%-1.94%), respectively (**Error! Reference source not found.**, **Error! Reference source not found.**, **Error! Reference source not found.**, **Error! Reference source not found.**). Dual-axis charts reveal that the highest number of dengue fever cases occurred in 2015, and the highest number of dengue-related deaths in 2017 within the scope of this study. The overall development trends for both are similar, yet there is a noticeable lag for the peak in mortality compared to the incidence. Notably, in terms of the impact of DALYs and deaths due to dengue fever, the values for males significantly exceed those for females, while the overall gender trend for incidence and prevalence is inverse (Figure 1). This disparity is more pronounced in the 2021 gender-stacked chart (Figure 2).

**Fig 1.**
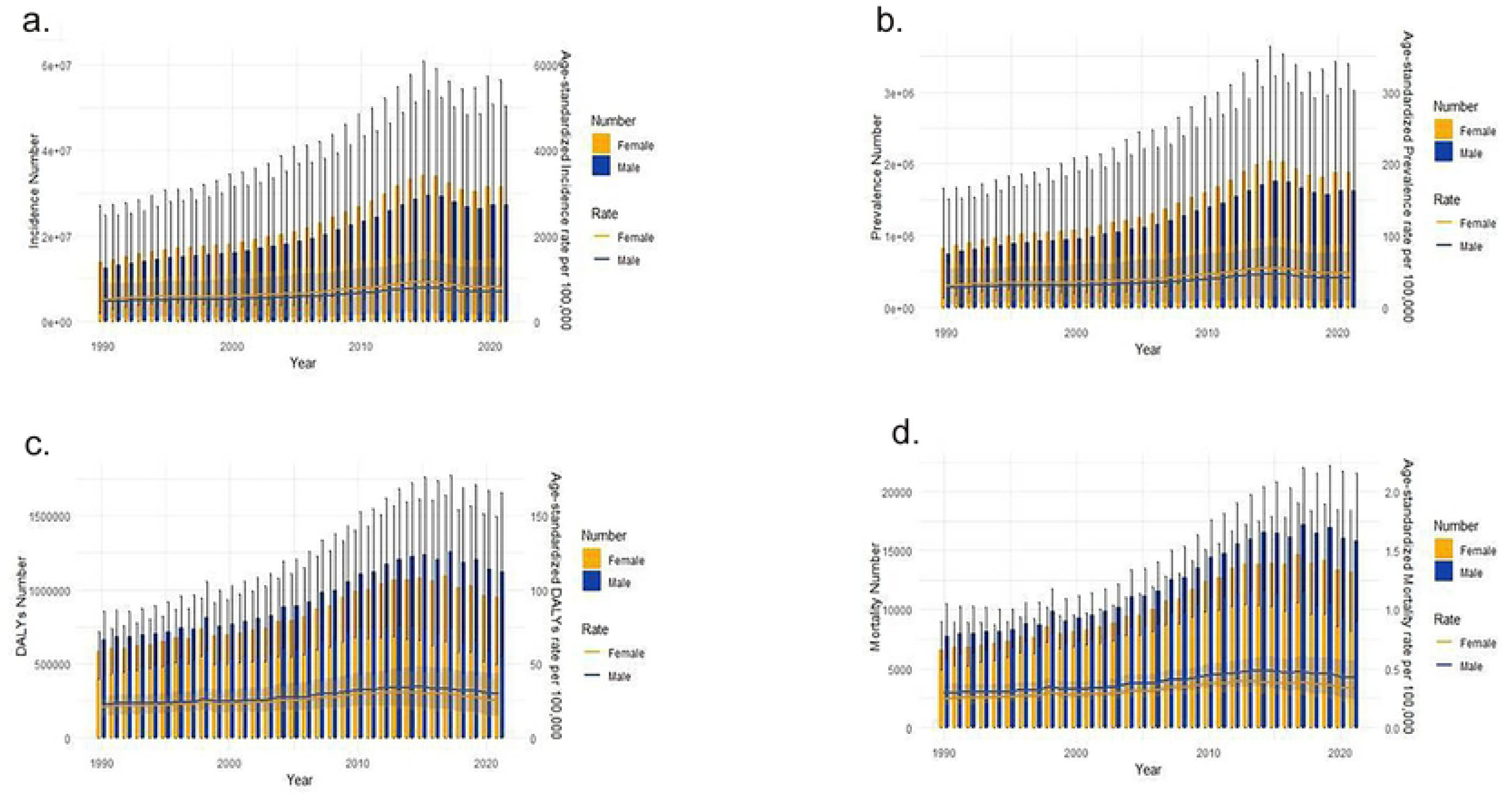
1990-2021 Dengue Trend. Fig 1 legend: The number and ASR of incidence (a), prevalence (b) DALYs(c), and mortality(d) in 1990-2021 Dengue Trend. Abbreviations: DALYs, disability-adjusted life-years.

**Fig 2.**
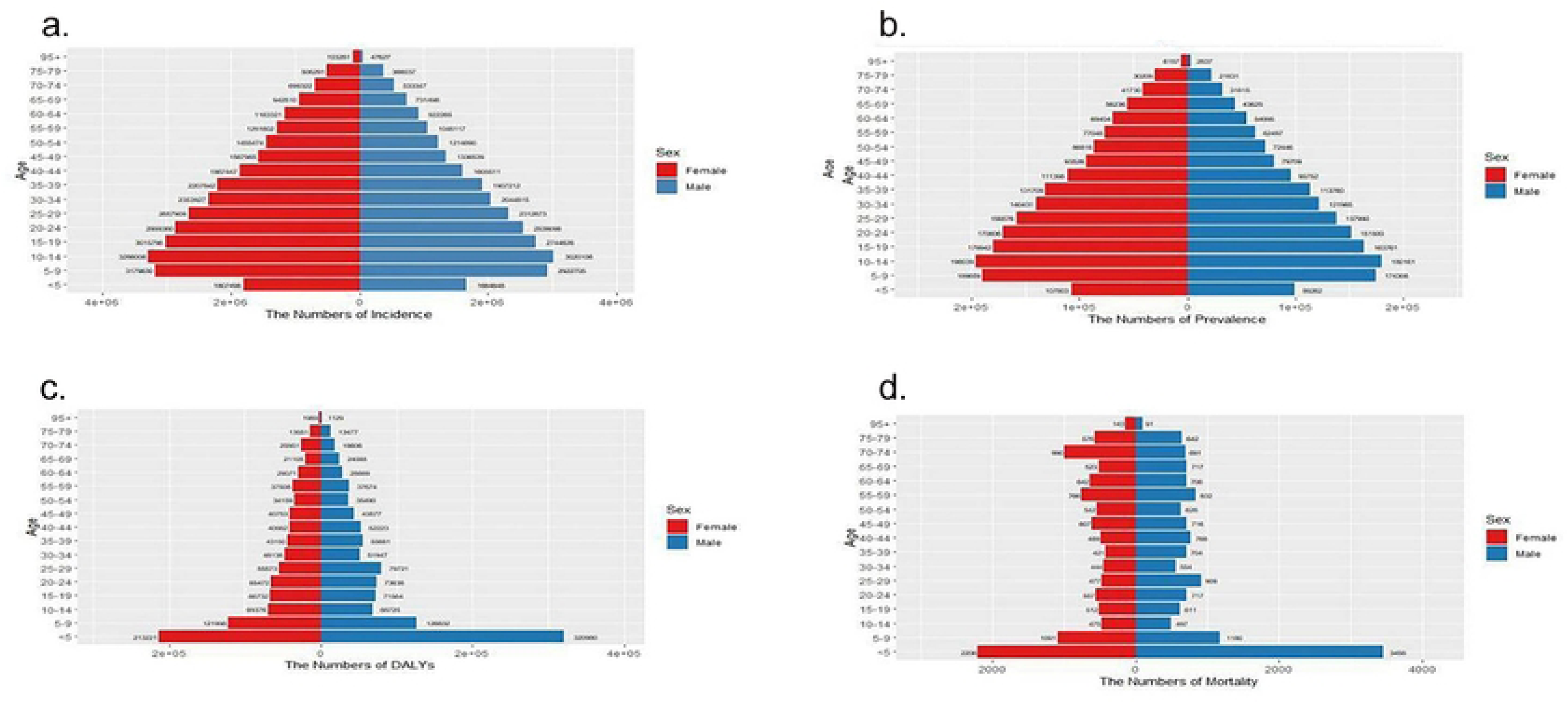
2021 Dengue’s gender-stacked chart. Fig 2 legend: The number of incidence(a), prevalence (b), DALYs (c) and deaths (d) of Dengue in Males and Females in 2021. Abbreviations: DALYs, disability-adjusted life-years.

At the regional level, the EAPC in the ASIR and ASPR are nearly congruent, with the high-income North America region showing the most significant increasing trend, while the Eastern Sub-Saharan Africa has the fastest decline. In the ASR for DALYs, the EAPC in the high-income North America region and Australia remains significantly elevated, but in the downward trend, Western Europe has taken the lead in place of Eastern Sub-Saharan Africa. Regarding the EAPC for the Age-Standardized Mortality Rate (ASMR), Tropical Latin America has the highest increasing trend, while Southern Latin America has the fastest decline (**Error! Reference source not found.**, **Error! Reference source not found.**, **Error! Reference source not found.**, **Error! Reference source not found.**).

At the national level, the overall trends in disease burden vary. Among various countries and regions (Figure 3), the EAPC in the ASR for both incidence and prevalence was highest in Tonga and lowest in South Sudan. However, in the EAPC for the age-standardized Disability-Adjusted Life Years rate (ASDR), Equatorial Guinea exhibited the highest increasing trend, while South Sudan had the most significant decreasing trend. Surprisingly, in the EAPC for the ASMR, Paraguay had the highest increasing trend, and Kuwait was at the forefront of the decreasing trend.

**Fig 3.**
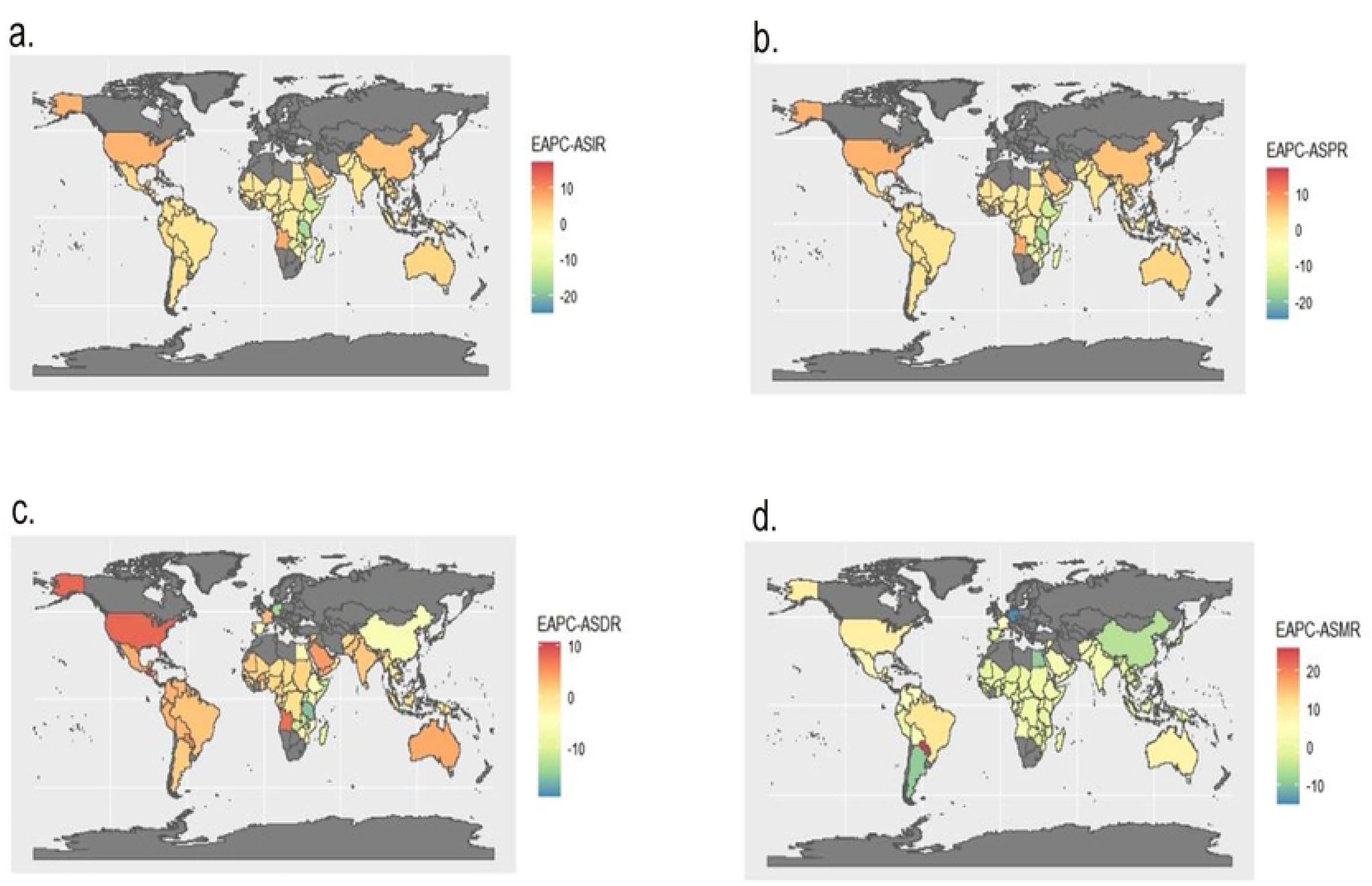
The EAPC trends in ASR for incidence, prevalence, DALYs and mortality. Fig 3 legend: (a) The EAPC Trends in ASIR of Dengue from 1990 to 2021; (b) The EAPC Trends in ASPR of Dengue from 1990 to 2021; (c) The EAPC Trends in ASDR of Dengue from 1990 to 2021; (d) The EAPC Trends in ASMR of Dengue from 1990 to 2021. Abbreviations: ASIR, age-standardized incidence rate; ASPR, age-standardized prevalence rate; ASDR, age-standardized disability-adjusted life-years rate; ASMR, age-standardized mortality rate; EAPC, estimated annual percentage change.

Additionally, when examining the range of SDI quintiles, the EAPC in the ASIR and ASPR for regions in the high-middle SDI quintile showed the most significant increasing trend among all SDI ranges; conversely, the EAPC for ASIR and ASPR in the low SDI quintile exhibited the fastest decline, with both trends being nearly congruent. Regarding the EAPC changes in the ASDR and ASMR due to dengue fever, regions with high SDI experienced the fastest rate of increase. However, in the EAPC for ASDR, regions with low SDI also showed an increasing trend, albeit at the slowest rate. The EAPC for ASMR showed the least change in regions with high-middle SDI (Tables S1, Table S2, Table S3, Table S4).

### Age-period-cohort analysis on Dengue incidence and prevalence

The results of age-period-cohort analysis for the incidence and mortality of dengue fever are presented in Figures 4 and 5.

**Fig 4.**
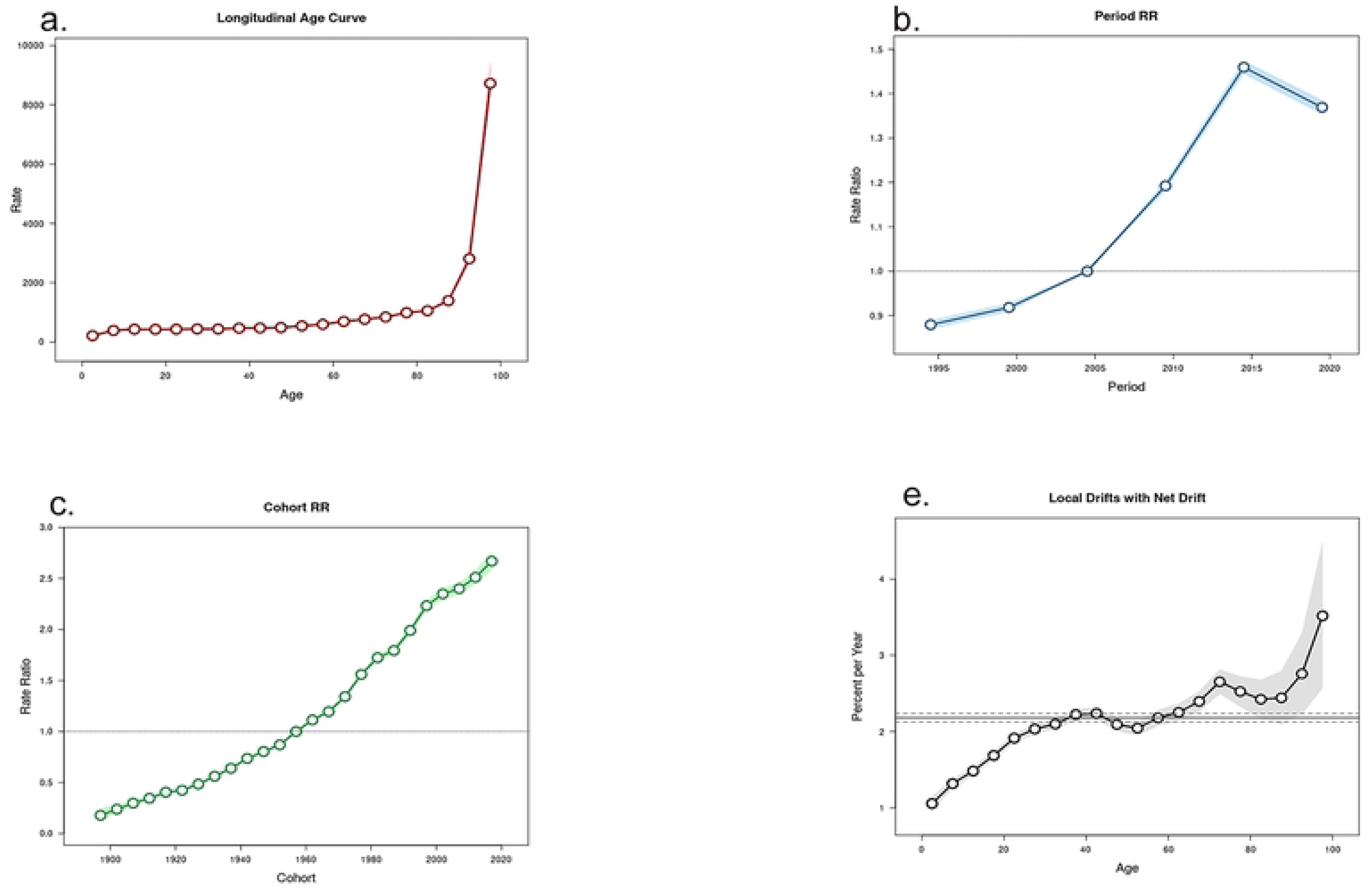
Dengue’s incidence age-period-cohort model (1) Fig 4 legend: Longtidunal age curve (a); period relative rate (b); cohort relative rate (c); local drifts with net drift (d); of incidence. Abbreviations: RR, relative risk.

**Fig 5.**
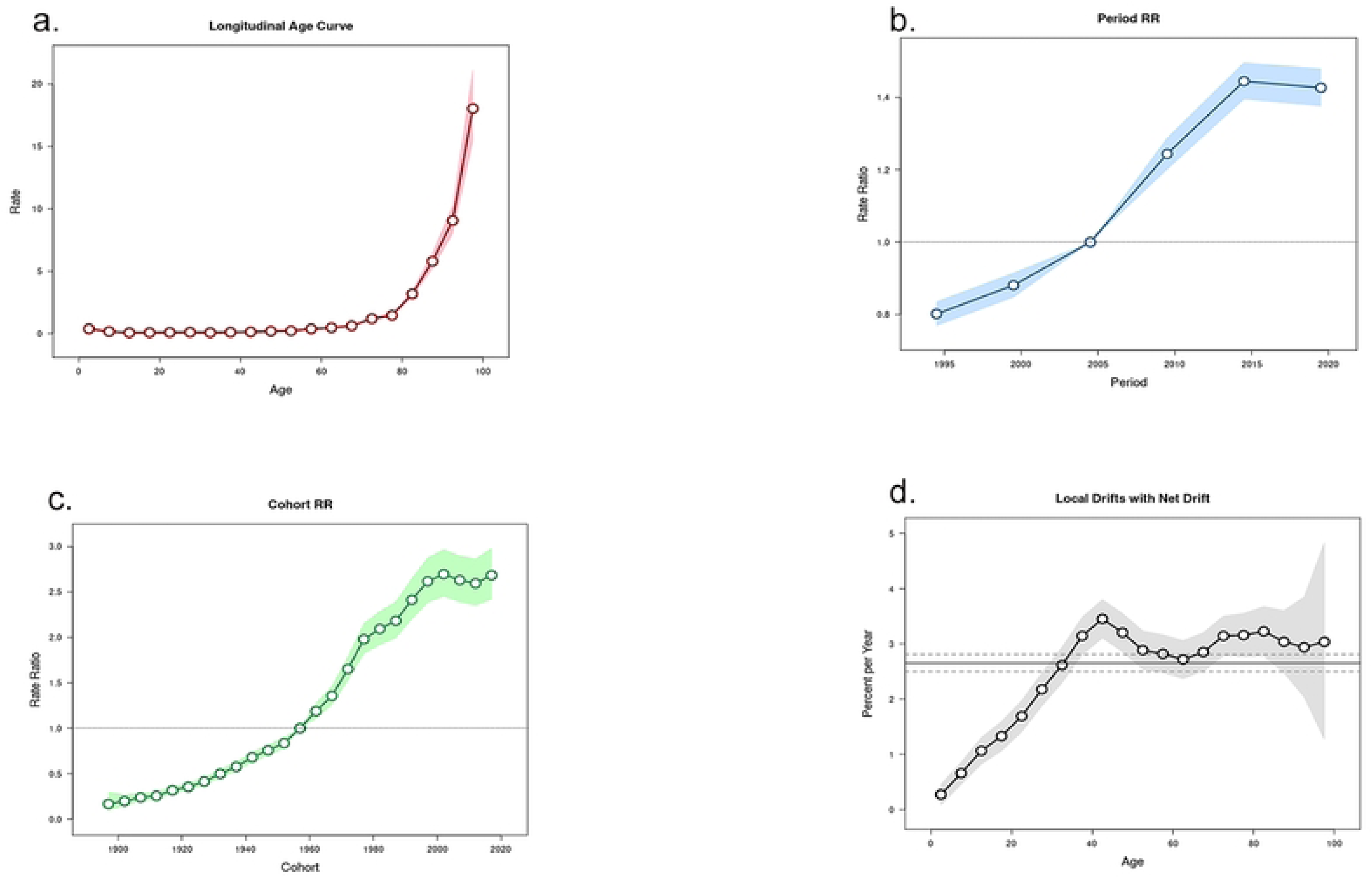
Dengue’s mortality age-period-cohort model (2) Fig 5 legend: Longtidunal age curve (a); period relative rate (b); cohort relative rate (c); local drifts with net drift (d); of mortality. Abbreviations: RR, relative risk.

After controlling for period and birth cohort effects, the age effect was shown to significantly influence the incidence and mortality of dengue fever, with both being positively correlated with age. Among those under 80 years old, the impact of age on the prevalence and mortality of dengue fever was relatively flat, while in individuals over 80 years old, the incidence and mortality rates of dengue fever increased linearly.

After controlling for age and birth cohort effects, the period effect had a significant impact on the incidence and mortality of dengue fever. The model used the incidence rate of dengue fever in 2005 as the baseline level, showing a long-term increasing trend from 2005 to 2015, with a relative risk (RR) increase of approximately 1.5 times compared to 1990, and serving as an inflection point for the subsequent decline in RR values. The mortality rate of dengue fever consistently showed an increasing trend from 1990 to 2015, and it was not until after 2015 that a decline in mortality rates began to emerge.

After controlling for the effects of age and period, the birth cohort effect significantly influences the risk of incidence and mortality of dengue fever. The birth cohort effect suggests that compared to earlier cohorts, individuals born between 2015 and 2019 have the highest risk of developing the disease. Moreover, as time progresses, the relationship between the RR and time approximates a directly proportional function trend.

It is noteworthy to compare the incidence of dengue fever across different time periods within the same age groups. The results indicate a sustained increase in the incidence of dengue fever among individuals aged 0-40, while those aged 40-45 continue to show an upward trend, albeit at a stable rate. A turning point is observed in the upward trend among individuals over the age of 70, with a subsequent sharp increase in the incidence of dengue fever in those over 90. Mortality rates, compared to the same point in the previous year, exhibit an almost directly proportional increasing trend in individuals aged 0-45. For those aged 45-60, the rate of increase slows down relative to the baseline level. However, for individuals over 60, mortality rates continue to fluctuate upwards, with an overall trend of continuous increase (Table S5 and S6).

### Decomposition analysis on Dengue DALYs

Over the past 32 years, there has been a sharp global increase in DALYs, with varying impacts of aging, population growth, and epidemiological changes across different SDI quintiles. Notably, the highest increase was observed in the middle SDI quintile. As of 2021, global aging contributed a negative growth of 29.55%, while population and epidemiological changes accounted for 74.45% and 54.80% of the global increase, respectively. The most significant contributions of aging, population growth, and epidemiological changes were noted in the middle SDI quintile with a negative growth of 58.33%, a growth of 92.41% in the low SDI quintile, and a growth of 85.08% in the middle SDI quintile, respectively (Table S7). When stratified by gender, the impacts of aging, demography, and epidemiology on DALYs varied across subgroups, with the burden being higher in males across all subgroups. The influence of population and epidemiology on DALYs was most pronounced in all SDI quintiles across all subgroups. Interestingly, the negative impact of aging was most pronounced in the middle SDI quintile, population growth had a significant impact on low-middle SDI regions, and the changes in DALYs due to dengue fever did not follow a simple ranking according to the definition of the quintiles from high to low (Figure 6).

**Fig 6.**
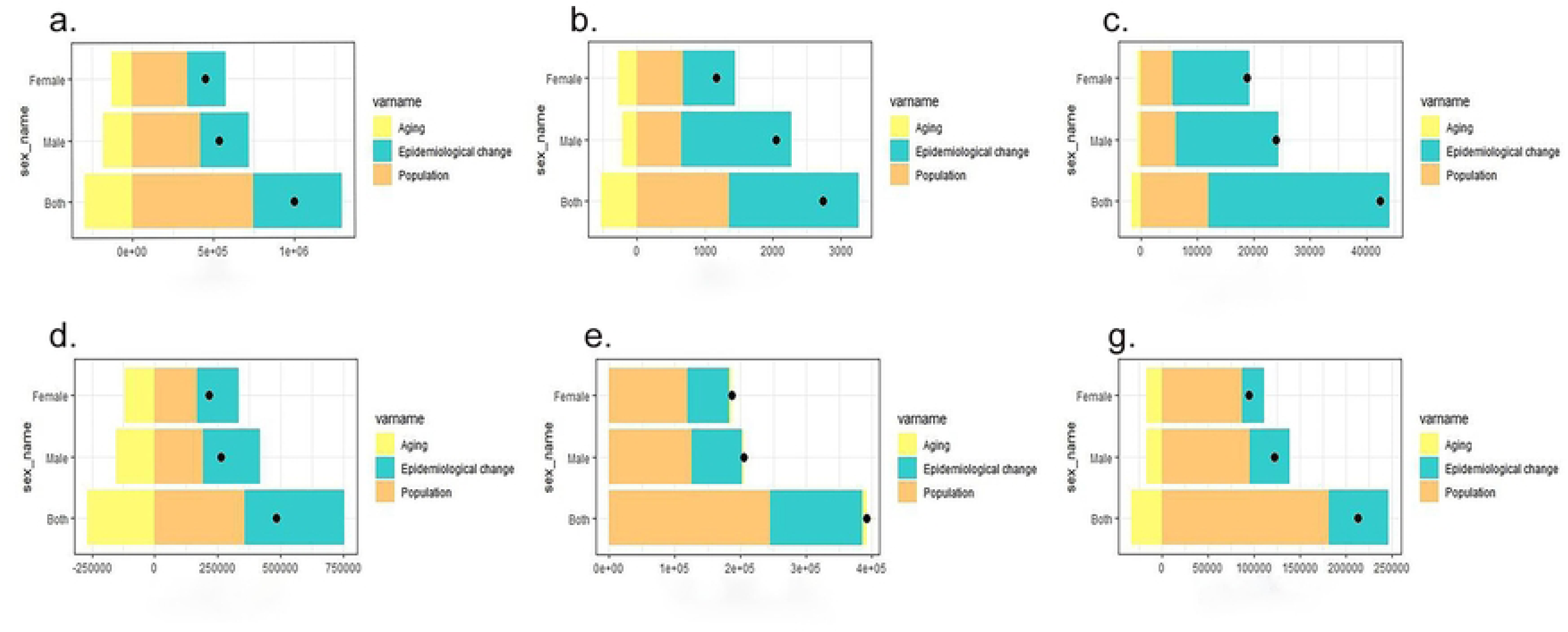
Decomposition analysis on dengue DALYs. Fig 6. Changes in DALYs of Dengue according to aging, population growth and epidemiological change from 1990 to 2021 at global level by SDI quintile and by subgroups of sexes. The black dot denotes the overall value of the changing resulting from all three components. For each component, the magnitude of a positive value suggests a corresponding increase in Dengue DALYs attributed to the component; the magnitude of a negative value suggests a corresponding decrease in Dengue DALYs attributed to the component. Abbreviations: DALYs, disability-adjusted life-years; SDI, sociodemographic index.

### Cross-country inequality analysis

Absolute inequalities in the burden of dengue fever in relation to the SDI were observed, with the slope index of inequality for 2021 being 1.29 (95% CI: −0.76 to 3.34), compared to −3.70 (95% CI: −4.94 to −2.46) in 1990. The impact of the SDI on the disease burden of dengue fever has diminished, with greater equality in 2021. Over time, from 1990 to 2014, the inequality in the burden of dengue fever, as measured by the number of reported cases worldwide, gradually decreased and even tended toward equality. However, during the subsequent period from 2015 to 2021, inequality intensified again, and the beneficiary groups shifted from subgroups in poorer groups to those in wealthier groups. Concurrently, the concentration index, which measures relative gradient inequality, was 0.26 (95% CI: 0.21-0.29) in 1990 and, when taken as an absolute value in 2021, was 0.15 (95% CI: 0.13 to 0.17). This indicates an improvement in the relative inequality of the burden of dengue fever, with a clear shift in the advantaged groups, namely from the poor in 1990 to the wealthy in 2021 (Figure 7).

**Fig 7.**
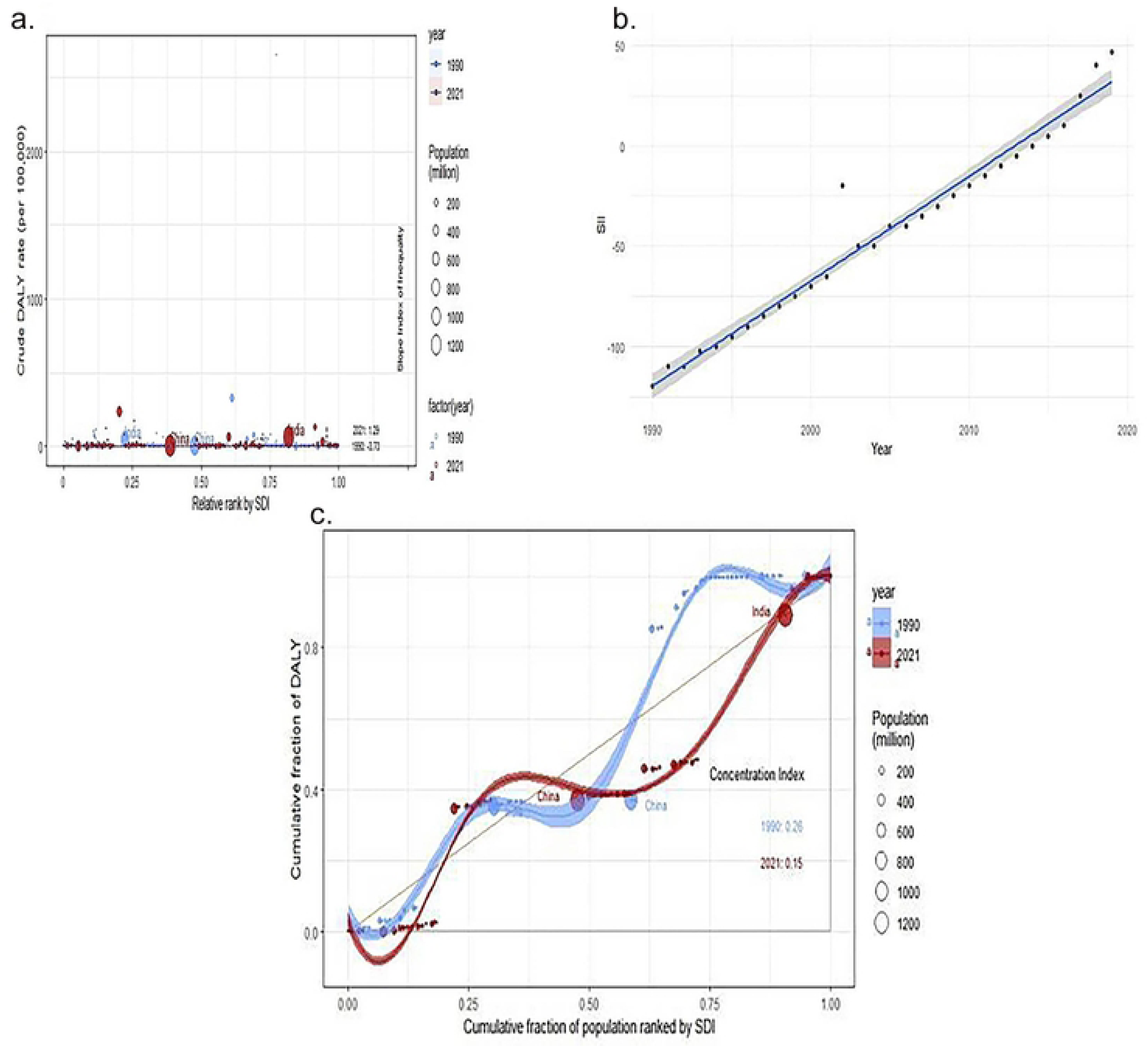
Analysis of cross-country inequality health. Fig 7. SDI-related health inequality regression for the DALYs of Dengue worldwide,1990 and 2021 (a); Trends in the SII for Dengue from 1990 to 2021 (b); concentration for the DALYs of Dengue worldwide,1990 and 2021 (c). Abbreviations: DALYs, disability-adjusted life-years; SDI, sociodemographic index, SII, Spatiotemporal Index of Infection.

### Predictive analysis on Dengue burden to 2051

The ASR for incidence, prevalence, DALYs, and mortality of dengue fever projected to 2051 are depicted in Figure 8. Globally, the ASR for incidence and prevalence is anticipated to stabilize, while the ASR for DALYs and mortality is projected to continue declining through 2051, with a more pronounced decline observed in females compared to males. Detailed values can be found in Table S8.

**Fig 8.**
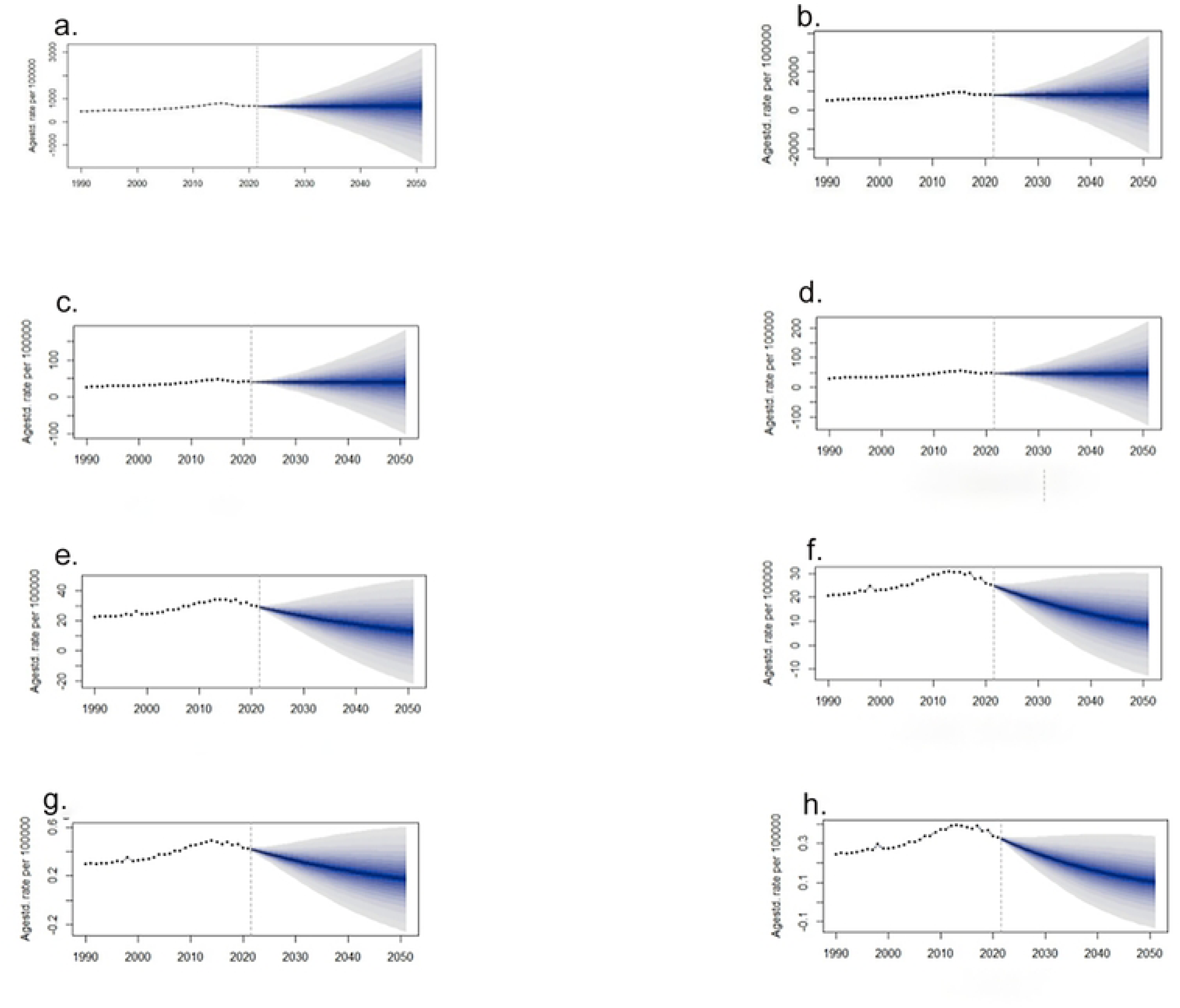
Predictive analysis on dengue burden. Fig 8. (a) The predicated ASIR for males from to 2051; (b) The predicated ASIR for females from to 2051; (c) The predicated ASPR for males from to 2051; (d) The predicated ASPR for females from to 2051; (e) The predicated ASDR for males from to 2051; (f) The predicated ASDR for females from to 2051; (g) The predicated ASMR for males from to 2051; (h) The predicated ASMR for females from to 2051; of Dengue globally. Abbreviations: ASIR, age-standardized incidence rate; ASPR, age-standardized prevalence rate; ASDR, age-standardized disability-adjusted life-years rate; ASMR, age-standardized mortality rate; DALYs, disability-adjusted life-years.

## Discussion

Through the comparative analysis presented, this study provides a detailed examination of the global, regional, and national burden of dengue fever from 1990 to 2021, based on trends, decomposition, health inequalities, and predictive analysis. Although there are differences in incidence, prevalence, DALYs, and mortality rates across countries, the global burden of dengue fever has generally shown an increasing trend from 1990 to 2021, with a shift in the beneficiary groups of health inequality during this period.

In terms of SDI quintiles, the disease burden of dengue fever is no longer solely a concern for third-world countries; developed nations such as the United States and Australia also face significant disease burdens due to dengue fever.

From a GBD regional perspective, compared to previous research on dengue fever [33,34], firstly, North America has become a hotspot for the spread of dengue fever. However, North America is not a geographically specific region for dengue fever transmission; the extensive spread of dengue fever in this area is often related to population movements, travel, and trade between countries [35,36]. Secondly, Tropical Latin America has maintained the highest dengue fever mortality rate globally over the past decades, which may be attributed to the region’s susceptibility to El Niño events, inconsistent governmental attitudes towards vaccination, and a lack of community-based mosquito control due to the prevalence of “urban diseases” [37]. Lastly, Western Europe and Southern Latin America have seen a decline in mortality rates due to dengue fever, likely associated with the unprecedented global attention to dengue fever in recent years. Notably, the World Health Organization(WHO)’s “Non-Communicable Disease Roadmap” and the “London Declaration” have made significant contributions to this trend [38,39].

Nationally, it is imperative to devise flexible health policies tailored to the specific circumstances of each country. For instance, Bali, a tourist destination in Indonesia, may not present travelers with a noticeable risk of mosquito bites during their visit, but many often contract dengue fever symptoms after returning from their trip. For Indonesia, effective surveillance and control of the disease among the traveling population is an urgent priority. Concurrently, it is noteworthy that the United States, as a leading economic power, has experienced an annual change in dengue fever incidence that has surpassed that of China. The focus of dengue fever prevention and control in the U.S. appears to be primarily on the 9-45 age group with a history of confirmed dengue infection, yet other high-risk age groups susceptible to the disease should be given particular attention through vaccination.

In the existing literature, there is a relative scarcity of studies that conduct a comprehensive analysis of the full age range of incidence and mortality data for dengue fever [40]. This study employs an age-period-cohort model for analysis and finds that individuals aged 60 and above have become a high-risk group for dengue fever infection and mortality. This phenomenon may be associated with the prevalence of chronic diseases, malnutrition, and decreased immunity among the elderly, all of which can exacerbate the disease burden. Therefore, special attention should be given to this group in disease prevention and control strategies [41,42,43]. Additionally, the bilateral chart analysis of this study for the year 2021 reveals that the number of dengue fever deaths among children and infants under the age of 10 remains high. This may be related to complex socio-political factors such as warfare, which could lead to a sharp increase in dengue fever mortality in this age group. Consequently, public health intervention measures should also give special consideration to children in this age range.

As the global population ages and continues to grow, epidemiological shifts occur rapidly, impacting different countries and regions in varying ways based on their economic levels and population dynamics. In regions with higher economic development, better access to public health resources leads to a reduced impact from population-related factors. At this juncture, without external policy interventions that highlight health equity, the disparities in health equity caused by epidemiological changes between impoverished and affluent regions will widen. Particularly for regions with high SDI and middle SDI, the disease burden of dengue fever exacerbated by population aging will further increase.

More importantly, the beneficiary groups of health equity may also shift due to policy differences between regions. Countries with high SDI levels have greater access to health care systems and health care systems perform better, thus potentially incurring a lower burden of disease[44]. According to this study, the health gap between the poorest and richest groups of dengue fever remains large, and the beneficiary group has shifted from poor to affluent groups. To promote health equity, it is imperative to bridge the vaccine divide in the international community. For many years, China and India, as the world’s most populous countries, have been frequently affected by dengue outbreaks[45,46,47,48,49]. The two have some valuable experience in the prevention and control of dengue fever, such as the release of “technical mosquitoes” for mosquitoes to use contraception to reduce transmission[50,51]; reduce carbon emissions and actively respond to the United Nations Framework Convention on Climate Change (UNFCCC)[52]; strengthen multimedia public welfare education to raise public awareness and community participation; placing mosquito killer lamps in public areas, continuous monitoring of cases in the community, and actively seeking international cooperation[53,54].

Although current predictive models indicate a potential decline in the ASR of DALYs and mortality for dengue fever by 2051, our study’s model provides a range of values that account for the multifactorial influences on dengue incidence and reporting. The potential for numerous factors to exacerbate the situation underscores the uncertainty in projecting the disease burden of dengue fever over the coming three decades. The incidence and reporting of dengue fever is still influenced by multiple factors. During the COVID-19 pandemic, for example, the number of cases reported to WHO decreased during this period. As a result, in 2023, another surge in dengue cases was observed worldwide. It is characterized by an increase in number and scale and the simultaneous occurrence of multiple outbreaks that have spread to areas previously unaffected by dengue. In addition, most patients with this disease have primary asymptomatic infection, and most of them do not develop clinical symptoms until the secondary infection. At the same time, dengue reporting is not mandatory in many countries[55].

Therefore, it is important to be aware that the incidence of dengue fever is likely to continue to rise as the world’s population continues to grow, with advances in surveillance and diagnostic technology[55]. The WHO and governments should pay close attention to the incidence of dengue fever in high-risk age groups and areas with high epidemic density, and actively work to improve health care systems in poor countries. At the same time, WHO should make full use of sociodemographic data and epidemiological changes to focus on the impact of accelerated population ageing on the increased burden of disease in countries around the world. In addition, in the context of the shift of beneficiaries of health inequality to affluent populations, WHO should once again carefully consider the rational allocation of medical resources and actively provide assistance to developing countries, and at the same time call on countries to strengthen public health education on dengue prevention and control, and work with other countries to respond to the UNFCCC and work together to address the impact of climate change on global health[52].

## Conclusion

In conclusion, dengue’s escalating cases and uneven global spread pose significant challenges for disease control. This study’s insights could aid in crafting tailored, adaptable public health policies and resource allocation, crucial for policymakers to enhance personalized healthcare systems.

## Data Availability

All relevant data are within the manuscript and its Supporting Information files.

## Funding

This work was supported by the National Natural Science Foundation of China (NSFC) funded project [82104135].

## Author contributions

Especially, the first author (Mingzhu Zhou) and the co-first author (Yong He) have contributed equally to this work.

Conceptualization: Mingzhu Zhou.

Data curation: Mingzhu Zhou, Yong He.

Formal analysis: Liangmiao Wu.

Methodology: Mingzhu Zhou, Yong He.

Project administration: Mingzhu Zhou, Liangmiao Wu, Kaiyuan Weng.

Resources: Liangmiao Wu.

Supervision: Kaiyuan Weng.

Validation: Mingzhu Zhou, Yong He.

Visualization: Mingzhu Zhou.

Writing-original draft: Mingzhu Zhou.

Wrting-review & editing: Mingzhu Zhou, Yong He, Liaomiao Wu.

## Acknowledgements

We appreciate the works by the Global Burden of Diseases, Injuries, and Risk Factors Study GBD 2021 collaborators.

## Support information

Additional supporting information can be found in the supplementary Material section.

**Table S1. The case number and ASR of incidence of Dengue in 1990 and 2021 for both sexes by SDI quintiles and by GBD regions.**

**Abbreviations:** ASR, age-standardized rate; EAPC, estimated annual percentage change; UIs, uncertainty intervals; CI, confidence interval.

**Table S2. The case number and ASR of prevalence of Dengue in 1990 and 2021 for both sexes by SDI quintiles and by GBD regions.**

Abbreviations: ASR, age-standardized rate; EAPC, estimated annual percentage change; UIs, uncertainty intervals; CI, confidence interval.

**Table S3. The case number and ASR of DALYs of Dengue in 1990 and 2021 for both sexes by SDI quintiles and by GBD regions.**

Abbreviations: ASR, age-standardized rate; EAPC, estimated annual percentage change; UIs, uncertainty intervals; CI, confidence interval.

**Table S4. The case number and ASR of deaths of Dengue in 1990 and 2021 for both sexes by SDI quintiles and by GBD regions.**

Abbreviations: ASR, age-standardized rate; EAPC, estimated annual percentage change; UIs, uncertainty intervals; CI, confidence interval.

**Table S5. RRs of Dengue incidence and mortality for both sexes due to age, period, and birth effects (1).**

Abbreviations: RR, relative risks; CI, confidence interval.

**Table S6. RRs of Dengue incidence and mortality for both sexes due to age, period, and birth effects (2).**

Abbreviations: RR, relative risks; CI, confidence interval.

**Table S7. Changes in DALYs of Dengue according to disease categories and population-level determinants from 1990 to 2021.**

Abbreviations: DALYs, disability-adjusted life-years; SDI, sociodemographic index.

**Table S8. The predicted ASR of incidence,prevalence,DALYs and mortality of Dengue from 2022 to 2051 globally.**

Abbreviations: ASR, age-standardized rate; DALYs, disability-adjusted life-years; CrI, credible interval.

## Notes

### Competing Interest Statement

The authors have declared no competing interest.

